# Contextualising Single-Arm Ophthalmology Trials With Real-World Data: An Emulated Target Trial Comparing The Efficacy of Interventions For Neovascular Age-Related Macular Degeneration

**DOI:** 10.1101/2020.08.10.20171538

**Authors:** Darren S. Thomas, Aaron Y. Lee, Philipp L. Müller, Roy Schwartz, Abraham Olvera-Barrios, Alasdair N. Warwick, Praveen J. Patel, Tjebo F.C. Heeren, Catherine Egan, Paul Taylor, Adnan Tufail, on behalf of the UK AMD EMR Users Group

**Affiliations:** Institute of Health Informatics, University College London (UCL), London, UK; Department of Ophthalmology, University of Washington, Seattle, WA, USA; Department of Ophthalmology, University of Bonn, Bonn, Germany; Moorfields Eye Hospital NHS Foundation Trust, London, UK; University College London Institute of Ophthalmology, London, UK; National Institute of Health Research Biomedical Research Centre at Moorfields Eye Hospital and UCL Institute Ophthalmology, London, UK; Institute of Cardiovascular Science, University College London (UCL), London, UK

**Author notes:** **Correspondence to** Adnan Tufail, Moorfields Eye Hospital NHS Trust, 162 City Road, London, EC1V 2PD, United Kingdom. PJP has received research funding from Bayer, UK, and has acted as a consultant for Bayer UK, Genentech, Novartis UK, and Roche. RS has received travel expenses from Allergan (AbbVie) and is a board member of Eye-Wise diagnostics. PT has received a grant from Novartis Pharmaceuticals. AT, PJP and CE received a proportion of their funding from the Department of Health’s NIHR Biomedical Research Centre for Ophthalmology at Moorfields Eye Hospital and UCL Institute of Ophthalmology. The views expressed in this publication are those of the authors and not necessarily those of the Department of Health. **Funding information** The ABC Trial was funded by the Special Trustees of Moorfields Eye Hospital. Novartis Pharmaceuticals awarded a grant to cover the costs for extracting and storing electronic health records, but were not involved in the study.

**Keywords:** Macular Degeneration, Electronic Health records, Propensity Score, Bevacizumab

## Abstract

Methods of causal inference have shown promise in replicating randomised trials using real-world data recorded by Electronic Health Records (EHRs). We herein emulated a target trial on the intention-to-treat efficacy of off-label bevacizumab (q6w) *pro re nata* relative to fixed-interval aflibercept (q8w) for improving week-54 visual acuity of eyes affected by neovascular age-related macular degeneration. The bevacizumab arm (n 65) was taken from the ABC randomised controlled trial. A total of 4,471 aflibercept-treated eyes aligning with the ABC trial eligibility were identified from EHRs and synthetic control arms were created by emulating randomisation conditional on age, sex, and baseline visual read via exact matching and propensity score methods. We undertook an inferiority analysis on mean difference at 54 weeks; outcomes regression on achieving a change in visual acuity of ≥ 15, ≥ 10, and ≤ −15 Early Treatment Diabetic Retinopathy Letters (ETDRS) letters at week 54; and a time-to-event analysis on achieving a change in visual acuity of ≥ 15, ≥ 10, and ≤ −15 ETDRS letters by week 54. Our findings suggest off-label bevacizumab to be neither non-inferior nor superior to licensed aflibercept. While being no substitute for randomised controlled trials, emulated target trials could aid the interpretation of single-armed trials.

## # INTRODUCTION

The treatment of retinal disorders with inhibitors of vascular endothelial growth factor (VEGF) amounts to 12% of total Medicare expenditure and requires considerable capacity to meet the demands of outpatient attendances ^1^. Development of new therapies for retinal vascular disorders, too, is both costly and inefficient in part due to the time taken to recruit sufficient patients for both the active and control trial arms. Roughly one-in-four ophthalmology trials are single-armed ^2^, but without a comparator arm making informed decisions to proceed to further phases of clinical trials and their design is more difficult. This limitation may be addressed by the use of an external synthetic control arm ^3,4^.

A target trial – the hypothetical trial tailored to the causal effect of interest – can be emulated by wholly observational design ^5^ or by augmenting an existing single-arm trial with a synthetic arm representing the counterfactual outcome of interest ^6^. More recently, advances in causal inference methods ^7-9^ have demonstrated concordance with trial reference standards ^6,10–12^, suggesting that contextualising single-armed trials with a synthetic arm is possible.

We herein emulated a target trial on the relative efficacy of intravitreal administration of two inhibitors of VEGF (off-label bevacizumab vs. licensed aflibercept) for improving visual acuity of eyes affected by neovascular age-related macular degeneration (nAMD) after 54 weeks of treatment. Our emulated target trial compared the bevacizumab arm (n 65) of the ABC trial ^13^ – a prospective, double-masked, randomised, controlled trial – with a synthetic control arm from 31,151 eyes receiving aflibercept during routine care as recorded by Electronic Health Records (EHRs). The choice of intervention and comparator is based on the lack of randomised controlled trials comparing the two therapies, despite off-label bevacizumab being the most commonly used drug to treat nAMD in the USA ^14^. The size of the ABC Trial bevacizumab arm is similar to that used in single-armed early-phase clinical trials. Our report follows the proposed emulated target trial framework ^15^.

## # METHODS

### ## Study design

An emulated target trial was undertaken comparing week 54 outcomes of eyes that, during the ABC trial, received bevacizumab (1.25 mg) *pro re nata* at six-week intervals (q6w) after three loading injections (i.e. the bevacizumab trial arm) with an external synthetic control arm of eyes that received aflibercept (2 mg) during routine care on the intention to treat at fixed eight-week intervals (q8w) reflective of the posology at launch of the therapy (i.e. the aflibercept synthetic arm). The target trial protocol, detailed in accordance with proposed guidelines ^15^, is outlined in Table S1.

### ## Informed Consent and Ethics

The ABC trial was approved by the ethics committee at each clinical site with all patients giving written informed consent to participate. The Caldicott data protection guardian at each EHR site gave signed permission for the de-identifiable EHRs to be extracted for analysis.

### ## Subjects

#### ### Bevacizumab trial arm

The ABC trial was a prospective, double-masked, multicentre randomised controlled trial undertaken in the United Kingdom during 2006-08. The method has previously been reported ^16^. Briefly, 131 patients were randomised 1:1 to receive in the study eye: I) 1.25 mg bevacizumab intravitreally at q6w *pro ne rata* until week 48 after three mandatory loading injections at weeks 0, 6 and 12 (n 65) or II) the standard of care at the time (n 66); the latter arm being irrelevant to this study. Patients could receive 3–9 bevacizumab treatments (0–6 maintenance injections). Administration of injection at each visit was determined *pro ne rata* based on the presence of subretinal fluid, development of haemorrhage or classic choroidal neovascularization, loss of > 5 Early Treatment Diabetic Retinopathy Study (ETDRS) letters in association with new intraretinal fluid, or persistent intraretinal fluid ^17^. Patients underwent assessments for visual acuity at each visit before treatment was administered, with a final assessment at week 54. Best-corrected visual acuity measurements were assessed using ETDRS reading charts.

#### ### Aflibercept synthetic arm

Synthetic control arms were sampled from EHRs recording ophthalmologic care across 27 sites in England during 2012–18 (Medisoft Ophthalmology platform (http://www.medisoft.co.uk), Leeds, UK). Medisoft EHRs hold anonymised longitudinal data on all episodes of ophthalmologic care from treatments & surgeries through to visual acuity measurements, diagnostics and complications. All visual acuity measurements were the best-recorded unaided and were measured via, or converted to, ETDRS letters. Qualitative assessments of counting fingers, hand motion, light perception, and no light perception were converted to ETDRS letter scores of 2, 0, 0, and 0, respectively.

Study eyes were the first diagnosed (or randomly sampled for bilateral diagnosis on same day) that initiated treatment with aflibercept (2 mg q8w fixed-dose after three loading injections at approximately four-weekly intervals within 70 days) for nAMD (for EHR phenotype, see Table S2). The pool of potential synthetic controls were eyes that would, as far as we could judge, have been eligible for the ABC trial using the eligibility criteria in Table S1 (for criteria not recorded by EHRs, see Table S3). In addition to the ABC eligibility criteria, we also excluded all eyes having an incomplete loading phase; all those that had unrecorded sex, age, or baseline visual acuity required for matching or propensity scoring; and all eyes that initiated treatment ≥ 1 December 2017 (allowing for 54-weeks follow-up).

### ## Emulating randomisation

Three methods (exact matching (EM), inverse probability treatment weighting (IPTW), and propensity score matching (PSM)) for achieving conditional exchangeability were compared with an unconditional (UC) design wherein all eyes eligible were analysed post protocol alignment but without addressing exchangeability. Hypothesised confounders were age, sex, and visual acuity at baseline read ^18,19^. The study workflow is presented in Figure 1.

**Figure 1:**
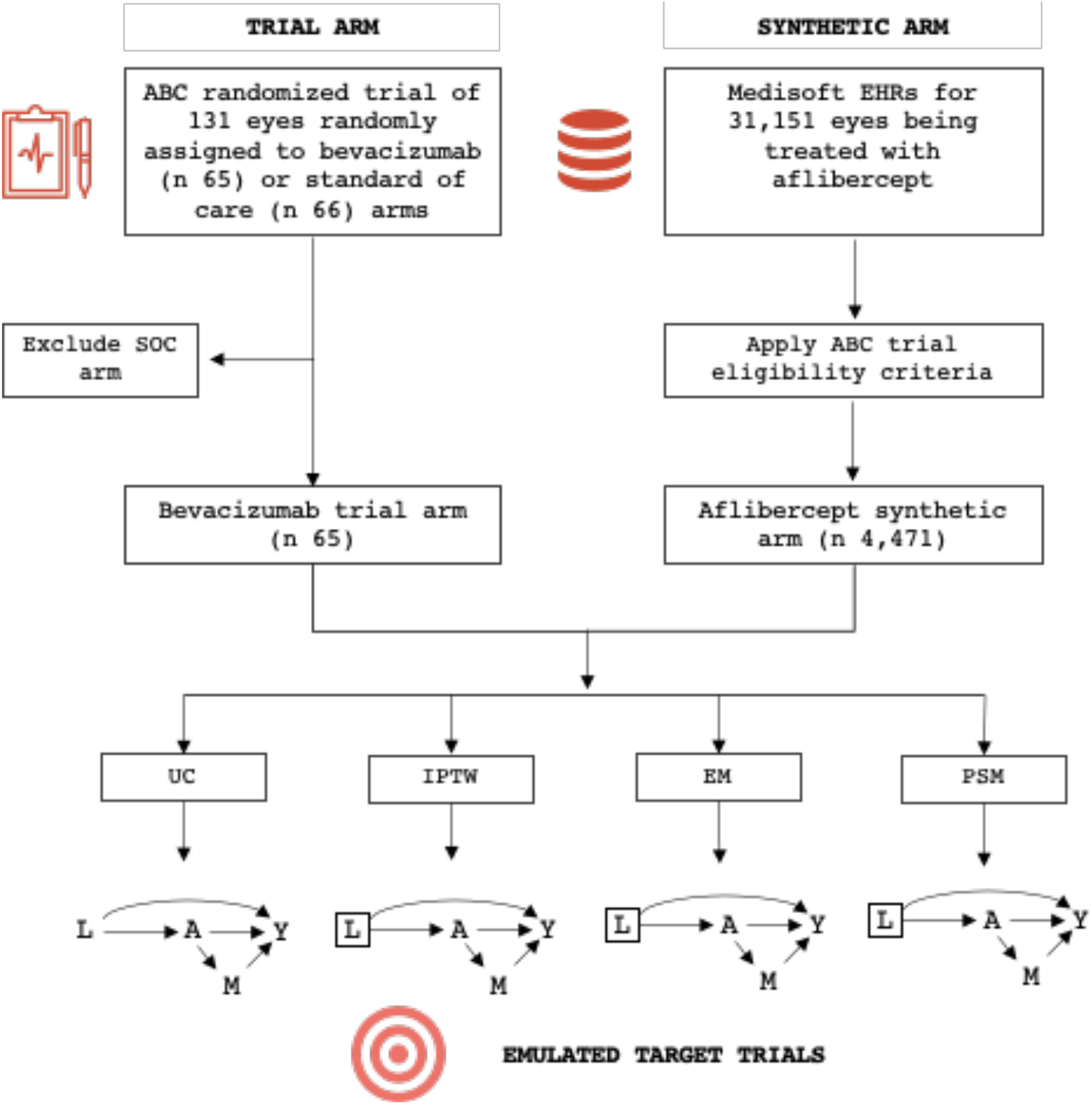
An emulated target trial estimating the causal effect of bevacizumab relative to aflibercept on the week-54 visual acuity of eyes affected by neovascular age-related macular degeneration. Age, sex, and ETDRS at baseline read (L) are common causes of both treatment assignment (A) and outcome (Y), while the number of maintenance injections received during the study period (M) mediates between A and Y. Thus, blocking the backdoor criterion L-Y via emulated randomisation (denoted by □), which is assumed to naturally arise in the bevacizumab trial arm through randomisation, permits unbiased estimates of the causal pathway A-Y. The backdoor criterion remains open under UC. Abbreviations: EHRs, Electronic Health Records; SOC, Standard of Care; UC, Unconditional; IPTW, Inverse Probability of Treatment Weighting; EM, Exact Matching; PSM, Propensity Score Matching.

#### ### Unconditional analysis

The UC analysis included all potential synthetic controls that aligned with ABC and emulated target trial-specific criteria analysed without emulated randomisation.

#### ### Exact matching

The aflibercept arm was exactly matched 1:1 with the bevacizumab trial arm on baseline confounders (age, sex, and visual acuity). Those with > 1 exact matches were paired with one randomly selected, while those with no exact matches were excluded from the analyses.

#### ### Inverse propensity score weighting & propensity score matching

The propensity score (Pr[A|L]) describes the probability of being in the ABC bevacizumab trial arm conditional on confounding variables (age, sex, and visual acuity). The score was derived from a binomial logistic model trained on all 131 ABC trial subjects (65 & 66 randomised to receive bevacizumab & standard of care, respectively (Equation 1)). The synthetic distribution tails not overlapping with the propensity scores of the bevacizumab arm were trimmed prior to weighting and matching.

The IPTW analysis included all 65 bevacizumab-treated eyes and all aflibercept-treated eyes that fulfilled the ABC trial and target trial eligibility criteria. Outcomes for each eye were thereafter weighted on the conditional inverse probability of treatment (1 / Pr[A|L] for the bevacizumab arm and 1 / (1 – Pr[A|L]) for the aflibercept arm). For PSM, synthetic controls were matched 1:1 to each bevacizumab eye (n 65) within a calliper of 0.1 standard deviations.

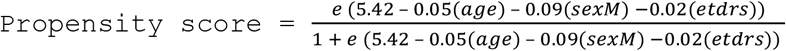

## Equation 1: Propensity score trained on ABC trial data

### ## Statistical analyses

We estimated intention-to-treat effects under each method for emulating randomisation, including all eyes that started treatment with bevacizumab or aflibercept regardless of the drug load received, treatment crossover or number of post-baseline measurements during the study period. Time zero/baseline for all analyses was on initiation of treatment. Baseline reads were taken on the day of first treatment or, to account for routine healthcare, the nearest to up to 30 days prior for EHR eyes. ABC eyes received their first treatment on the day of randomisation. Study exits were dependent on the analysis, as detailed later. Exchangeability in confounding variables was assessed pre and post emulating randomisation via standardised differences calculated as reported ^20^. A standardised difference < 0.1 was considered evidence for conditional exchangeability. Statistical inferences were made at an α threshold of ≤ 0.05.

#### ### Non-inferiority

An ordinary least squares regression was modelled on the change in visual acuity from baseline to week 54 measurement, stratified by treatment (aflibercept arm as reference). Non-inferiority was declared if the lower confidence interval bound was ≥ than a non-inferiority margin of –4 ETDRS letters. If no measurement was taken during week 54 owing to the inexactness of routine healthcare attendances, we used the measurement taken nearest to week 54 during weeks 50–58, taking the earliest date if two dates were equally tied. Missing data was imputed using the last observation carried forward.

#### ### Binomial outcomes

A binomial logistic regression, stratified by treatment, was modelled on the outcomes of achieving a change in visual acuity from baseline to week 54 measurement of ≥ 15 letters, ≥ 10 letters, and ≤ -15 letters. We present the Odds Ratios (ORs) and 95% confidence intervals (CIs) relative to the aflibercept synthetic arm as reference. As for the non-inferiority analysis, if no measurement was taken during week 54, we used the measurement taken nearest to week 54 during weeks 50–58, taking the earliest date if two dates were equally tied. Missing data was imputed using the last observation carried forward.

#### ### Time-to-event

Kaplan-Meier estimators were modelled on the time from baseline to achieve a change in visual acuity of ≥ 15 letters, ≥ 10 letters, and ≥ -15 letters, stratified by treatment (aflibercept arm as reference). Right censorship was on the last visual acuity measurement taken during the study period, up to 54 weeks maximum. Inferences were made on the outputs of the log-rank test comparing event distributions.

### ## Software

EHRs were extracted by Medisoft (Leeds, UK) and stored as relational tables on a MySQL server (version 5.5.62). All analyses were undertaken with R version 4.0.2^21^ with particular use of tidyverse version 1. 3.0^22^, survival version 3.2–3^23^, and fuzzyjoin version 0.1.6^24^. All code and an exhaustive listing of all R packages used is available at https://github.com/dsthom/synthetic_trial_amd.

## # RESULTS

### ## Subjects

A pool of 31,151 eyes that started aflibercept treatment for nAMD was identified. Of these, 26,680 (86% of 31,151) were ineligible, either not fulfilling the ABC (n 20,823 (67%)) or the emulated trial criteria (n 5,857 (19%)). Thus, a pool of 4,471 (14%) eyes were available for study. Of these, 186 eyes were trimmed from the IPTW and PSM analyses for having non-overlapping propensity scores for a pool of 4,141 (0.13) eyes. A stepwise breakdown of ineligibility is presented in Table S4. Eyes included in each bevacizumab trial & aflibercept synthetic arms were respectively, 65 & 4,471 for UC; 131 & 8,506 for IPTW (pseudo-population); 43 & 43 for EM; and 65 & 65 for PSM.

The aim of emulating randomisation was to achieve conditional exchangeability in measured confounding (summarised in Table 1). Prior to aligning protocols, arms were imbalanced on age and baseline ETDRS, and after aligning protocols (UC) on age only. IPTW and EM were balanced for all measured confounding, while imbalances remained post PSM for age and the process in itself seemed to imbalance baseline ETDRS. The causal assumption of positivity was satisfied for all levels of confounding.

**Table 1:** Baseline traits of emulated target trial arms.

| Variable | Aflibercept synthetic arm | Bevacizumab trial arm | Standardised difference |
| --- | --- | --- | --- |
| **Pre-aligning of trial protocols (n 25,199)** |  |  |  |
| Arm size | n 25,134<sup>1</sup> | 65 | — |
| Sex (Female) | n 15,860 (63%) | n 39 (60%) | 0.06 |
| Age in years | $\bar{x}$ 80 ($\sigma_x$ 8) | $\bar{x}$ 79 ($\sigma_x$ 8) | 0.14 |
| Baseline ETDRS | $\bar{x}$ 56 ($\sigma_x$ 17) | $\bar{x}$ 51 ($\sigma_x$ 12) | 0.34 |
| Propensity score | $\bar{x}$ 0.48 ($\sigma_x$ 0.12) | $\bar{x}$ 0.52 ($\sigma_x$ 0.10) | 0.34 |
| **Unconditional (n 4,536)** |  |  |  |
| Sample n | n 4,471 | n 65 | — |
| Sex (Female) | n 2,817 (63%) | n 39 (60%) | 0.06 |
| Age in years | $\bar{x}$ 80 ($\sigma_x$ 8) | $\bar{x}$ 79 ($\sigma_x$ 8) | 0.14 |
| Baseline ETDRS | $\bar{x}$ 52 ($\sigma_x$ 13) | $\bar{x}$ 51 ($\sigma_x$ 12) | 0.09 |
| Propensity score | $\bar{x}$ 0.50 ($\sigma_x$ 0.11) | $\bar{x}$ 0.52 ($\sigma_x$ 0.10) | 0.17 |
| **Inverse Probability of Treatment Weighting (n 8,637<sup>2</sup>)** |  |  |  |
| Sample n | n 8,506<sup>2</sup> | n 131<sup>2</sup> | — |
| Sex (Female) | n 5,372 (63%) | n 81 (62%) | 0.03 |
| Age in years | $\bar{x}$ 80 ($\sigma_x$ 7) | $\bar{x}$ 80 ($\sigma_x$ 7) | 0.06 |
| Baseline ETDRS | $\bar{x}$ 52 ($\sigma_x$ 13) | $\bar{x}$ 52 ($\sigma_x$ 12) | 0.03 |
| Propensity score | $\bar{x}$ 0.50 ($\sigma_x$ 0.10) | $\bar{x}$ 0.49 ($\sigma_x$ 0.10) | 0.09 |
| **Exact matching (n 86)** |  |  |  |
| Sample n | n 43 | n 43 | — |
| Sex (Female) | n 29 (67%) | n 29 (67%) | 0 |
| Age in years | $\bar{x}$ 80 ($\sigma_x$ 6) | $\bar{x}$ 80 ($\sigma_x$ 6) | 0 |
| Baseline ETDRS | $\bar{x}$ 51 ($\sigma_x$ 11) | $\bar{x}$ 51 ($\sigma_x$ 11) | 0 |
| Propensity score | $\bar{x}$ 0.50 ($\sigma_x$ 0.09) | $\bar{x}$ 0.50 ($\sigma_x$ 0.09) | 0 |
| **Propensity Score Matching (n 130)** |  |  |  |
| Sample n | n 65 | n 65 | — |
| Sex (Female) | n 40 (62%) | n 39 (60%) | 0.03 |
| Age in years | $\bar{x}$ 78 ($\sigma_x$ 8) | $\bar{x}$ 79 ($\sigma_x$ 8) | 0.12 |
| Baseline ETDRS | $\bar{x}$ 53 ($\sigma_x$ 12) | $\bar{x}$ 51 ($\sigma_x$ 12) | 0.20 |
| Propensity score | $\bar{x}$ 0.52 ($\sigma_x$ 0.10) | $\bar{x}$ 0.52 ($\sigma_x$ 0.10) | 0 |
| <sup>1</sup> 31,151 minus eyes with unrecorded sex, age, or baseline ETDRS.<br><sup>2</sup> pseudo-population created by weighting. |  |  |  |

### ## Non-inferiority

Approval of novel nAMD therapies is often based on proof of their non-inferiority to currently approved drugs. Our results generally suggest, with exception of PSM, that bevacizumab is not inferior to aflibercept assessed on a noninferiority margin of 4 ETDRS letters (Figure 2). There was, however, insufficient evidence to conclude non-inferiority under PSM (−0.1 mean ETDRS difference relative to aflibercept). Confidence bounds for matching methods were notably wider than those for UC and IPTW.

**Figure 2:**
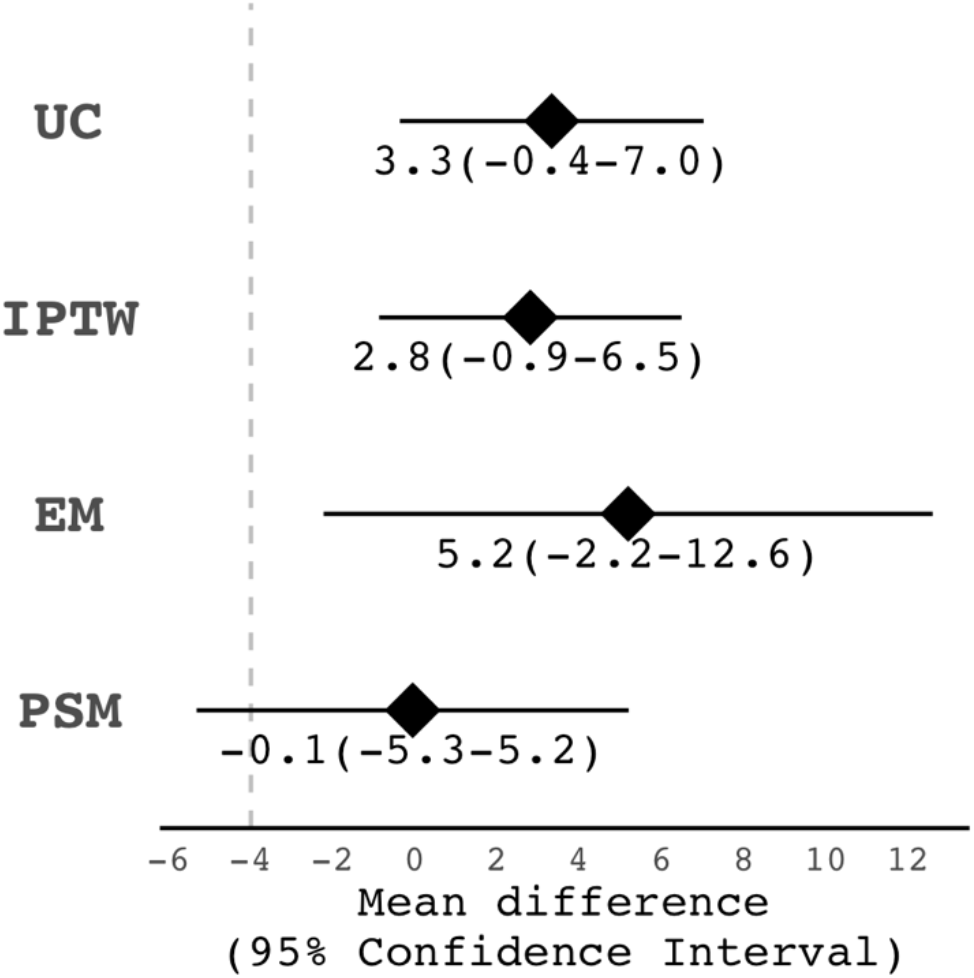
Mean ETDRS difference in change in visual acuity from baseline to week 54 measurement for bevacizumab-treated eyes relative to those treated with aflibercept. Dashed vertical line denotes a non-inferiority margin of -4 ETDRS letters. Abbreviations: UC, Unconditional; IPTW, Inverse Probability of Treatment Weighting; EM, Exact Matching; PSM, Propensity Score Matching.

### ## Superiority

We mirrored the primary analysis of the ABC trial in which the proportion of eyes in each treatment arm that gained ≥ 15 ETDRS letters at week 54 were compared (Figure 3). Secondary analysis on the proportion of eyes having a change in visual acuity from baseline to week 54 measurement of ≥ 10 letters and ≤ –15 letters are also presented. Under no trials tested was bevacizumab shown to be superior to aflibercept.

**Figure 3:**
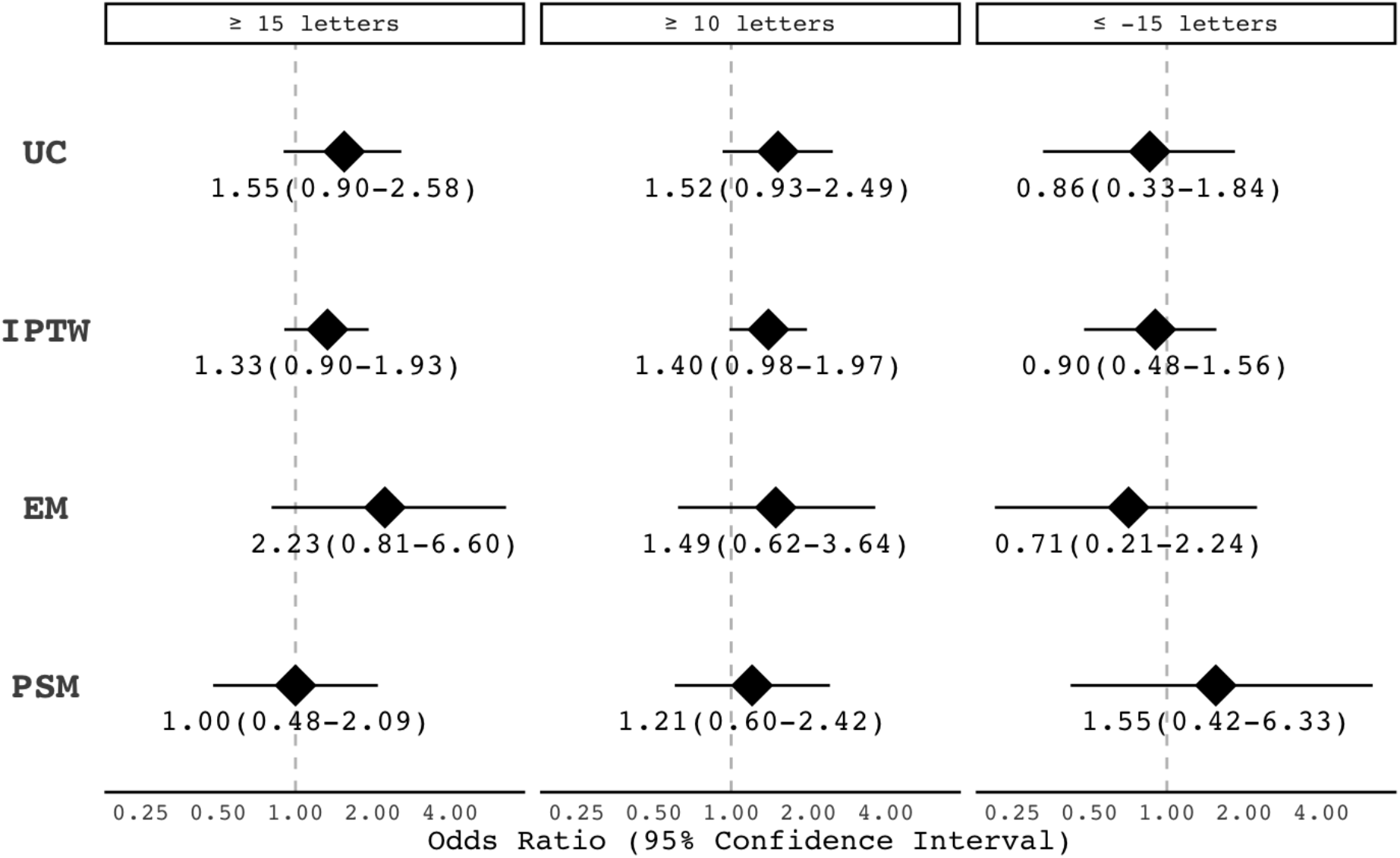
Odds of bevacizumab-treated eyes achieving *a priori* outcomes 3 at week 54 relative to those treated with aflibercept. Abbreviations: UC, Unconditional; IPTW, Inverse Probability of Treatment Weighting; EM, Exact Matching; PSM, Propensity Score Matching.

### ## Time-to-event

It may be that the drugs themselves, or the protocols under which they were administered, are beneficial at varying rates during the study period. We explored this possibility with the Kaplan-Meier estimator modelled on the time to a change in visual acuity from baseline of ≥ 15, ≥ 10 and ≤ -15 ETDRS letters (Figure 4). The log-rank statistic of event distributions suggests there being no difference in the event distribution of eyes receiving bevacizumab relative to aflibercept.

**Figure 4:**
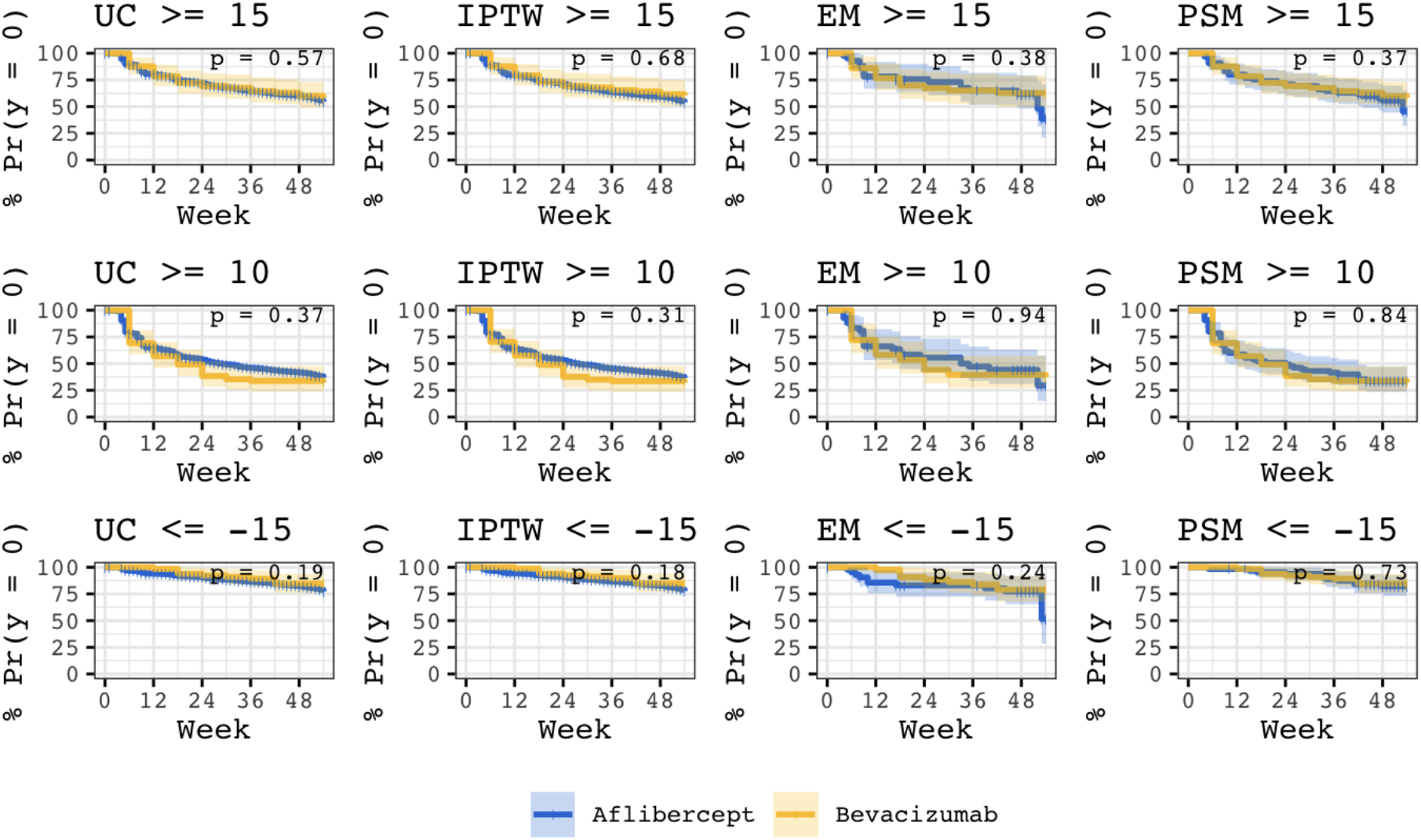
Hazards of bevacizumab-treated eyes achieving *a priori* outcomes during the first 54 weeks of treatment relative to those treated with aflibercept. Hued ribbons represent 95% confidence intervals. Abbreviations: UC, Unconditional; IPTW, Inverse Probability of Treatment Weighting; EM, Exact Matching; PSM, Propensity Score Matching.

The Kaplan-Meier curves, too, suggest disproportional hazards over time, though this may be an artefact of the fixed-interval measurements of the trial arm.

## # DISCUSSION

### ## Principal findings

Randomised controlled trials are the standard for causal inference but are not always practical, ethical, or timely to undertake. In answering an unaddressed clinical question, we herein emulated a target trial through use of causal inference methods to contextualise a single trial arm with an external synthetic arm derived from real-world data recorded by EHRs. Our findings suggest off-label bevacizumab to be neither non-inferior nor superior to licensed aflibercept for improving the week-54 visual acuity of eyes affected by nAMD.

### ## In context with the evidence base

Though we have no trial reference to validate the present findings with, we can attempt to contextualise ours in relation to the literature. At writing two randomized trials have reported on the efficacy of inhibitors of VEGF for treating nAMD; each having a common comparator arm, ranibizumab, from which we can triangulate. The CATT trial demonstrated bevacizumab to be non-inferior to ranibizumab ^25^.

The VIEW trial reported aflibercept to be non-inferior to ranibizumab^26^. These findings taken together suggests there being no biological reason to suspect bevacizumab to be inferior to aflibercept, which is consistent with the results of this emulated trial.

Secondly, the methods of causal inference used herein have been validated by a number of studies that assessed the extent to which a target trial can be emulated using real-world data. In a study on statins and cancer, the authors revisited the past failures of observational analogues to arrive at the findings of clinical trials.^8^ It was argued that this failure is not primarily due to confounding biases introduced by non-random treatment assignment, as is often thought, but also to biases introduced by subjects and analyses deviating from an explicit target trial protocol; advice that later evolved into the target trial framework ^15^ that we followed herein. Under the target trial approach, observational estimates were concordant with those of the trial that was emulated ^8^. In another study most applicable to ours is that in which the concordance in estimates obtained by emulated target trials composed of one trial arm and an external control arm derived from real-world data were compared with that of a reference two-armed trial ^6^. The paper reports, through alignment of protocols and propensity-based adjustment, concordance in ten out of the eleven trials emulated. It is on these validated methods that our findings are dependent on.

A criticism of PSM is that its use may inadvertently exacerbate exchangeability ^27^, and we question whether there is a link between this possibility and the discordance in the PSM non-inferiority estimate relative to other methods. Given the evidence base is that visual acuity on starting treatment is the most important determinant of visual improvement ^18, 28, 29^, it may therefore be plausible that the imbalance of the PSM aflibercept arm toward greater baseline acuity – despite exchangeability in the dimension of propensity score – nullified the relative efficacy of bevacizumab. Notably, of all methods for emulating randomisation, PSM was the only to lead to inexchangeability in baseline read, and more curiously was that this being exchangeable prior to emulating randomisation. Another notable limitation of matching in general is the reduced sample size leading to widened confidence bounds and potential biases from discarding unmatched observations. IPTW, in contrast, amplifies the weighting of these rare events without discarding data.

### ## Strengths & limitations

A major strength of our study is that the target trial we herein emulated has not yet been undertaken as a randomised controlled trial, and thus advances in our understanding of causal inference has enabled us to study these causal effects ethically, timely, and inexpensively.

Furthermore, the size and granularity of Medisoft EHRs enabled us to align our synthetic arm with the ABC trial protocol; and the few criteria it was not possible to mirror were minor and were unlikely to be influential.

Naturally, we also acknowledge several limitations. The first being the potential for residual confounding. Where trials can assume exchangeability across all confounders – measured and unmeasured – due to random treatment assignment, emulated target trials in contrast are limited to exchangeability only in measured variables. Secondly, the non-overlap in treatment periods between the bevacizumab (2006–08) and aflibercept (2012–18) arms could bias our estimates if the assumption of exchangeability does not hold. We have seen from our EHRs in the UK, for example, that eyes affected by nAMD have presented with greater visual acuity since 2008 (unpublished data), thereby implying that these eyes are more limited in the benefits treatment can provide. An ideal counterfactual arm would receive treatment concurrent to the single-armed trial.

### ## Implications for stakeholders

It is evident through the founding of the FDA Real-World Evidence Program ^30^ that regulatory decisions may increasingly rely on real-world evidence. Indeed, the use of external counterfactual arms has already expedited the regulatory approval of blinatumomab (Amgen) ^10^ and alectinib (Roche) ^12^ for oncological indications. Nevertheless, barring a small evidence base, the extent to which randomised trials can be emulated by real-world data is largely unknown. The FDA has also funded the RCT DUPLICATE initiative (rctduplicate.org), which aims to determine the validity of using real-world evidence for regulatory decisions ^4^. More immediately, our study has implications for the economics of treating nAMD for bevacizumab is somewhat more cost-effective than licensed therapies ^31^. For context, a 2018 survey by the *American Society of Retina Specialists* reports aflibercept being the first-line therapy for 13, 16, 37, 41, and 47% of ophthalmologists surveyed across Africa, the USA, Europe, Asia, and Central & South America ^14^.

### ## Unanswered questions & future research

With no existing trials comparing the efficacy of bevacizumab to aflibercept and with bevacizumab not being prescribed per ABC protocol during routine healthcare, it was not possible for us to triangulate our findings against a reference standard. We nevertheless acknowledge the importance of validating our findings, and aim to use the Medisoft EHRs to triangulate our methods relative to existing clinical trials. Furthermore, the estimation of per protocol effects, either by naive definition or by weighting on the conditional probability of treatment adherence ^32^, would be an important contribution to the evidence base. This would require the delineation of non-adherence from *pro re nata*.

## # CONCLUSIONS

Off-label bevacizumab administered *pro re nata* (q6w) is neither noninferior nor superior to licensed aflibercept administered at fixed intervals (q8w) for improving the week-54 vision of eyes affected by neovascular age-related macular degeneration.

## Data Availability

Data may be obtained from a third party but are
not publicly available. All code is hosted at https://github.com/dsthom/synthetic_trial_amd.

## # ACKNOWLEDGMENTS

We are ever thankful for the comments and suggestions of Dr Michail Katsoulis during the drafting of this manuscript.

## # AUTHOR CONTRIBUTIONS

AT, DST, PT, and AL conceived the idea and designed the research.

PT, AL, AT, and CE obtained the data.

DST undertook all analyses.

TFCH, ANW, and AO-B reviewed the code.

DST, AL, PLM, CE, PT, and AT wrote the first draft.

All authors contributed to the editing of the final manuscript.

## Notes

**Conflict of Interests** AL is an employee of the US FDA, has received research funding from Novartis, Santen, Carl Zeiss Meditec, and has acted as a consultant for Genentech, Topcon, and Verana Health. The opinions expressed in this article are the author’s own and do not reflect the view of the FDA or the United States government.

### Competing Interest Statement

AL is an employee of the US FDA, has received research funding from Novartis, Santen, Carl Zeiss Meditec, and has acted as a consultant for Genentech, Topcon, and Verana Health. The opinions expressed in this article are those of the author and do not reflect the view of the FDA or the United States government.
PJP has received research funding from Bayer, UK, and has acted as a consultant for Bayer UK, Genentech, Novartis UK, and Roche.
RS has received travel expenses from Allergan (AbbVie) and is a board member of Eye-Wise diagnostics.
PT has received a grant from Novartis Pharmaceuticals.
AT, PJP and CE received a proportion of their funding from the Department of Health NIHR Biomedical Research Centre for Ophthalmology at Moorfields Eye Hospital and UCL Institute of Ophthalmology. The views expressed in this publication are those of the authors and not necessarily those of the Department of Health.

